# Cost-effectiveness of diagnostic criteria for postpartum haemorrhage treatment

**DOI:** 10.1101/2025.09.02.25334895

**Authors:** Nick Scott, Ioannis Gallos, Thomas Walsh, Caitlin R Williams, John Allotey, Anderson Borovac-Pinheiro, Adam Devall, Mario R Festin, Shivaprasad S. Goudar, Christian Haslinger, G Justus Hofmeyr, Malcolm J Price, Zahida P Qureshi, Loïc Sentilhes, Idnan Yunas, Arri Coomarasamy, Olufemi T Oladapo

## Abstract

**Background:** A threshold of ≥ 500 ml blood loss within 24 hours of childbirth has conventionally been used to initiate postpartum haemorrhage (PPH) treatment. We assessed the cost-effectiveness of initiating PPH treatment at lower blood loss thresholds, alone and in combination with any other abnormal haemodynamic marker (pulse, systolic and diastolic blood pressure, shock index), compared with the conventional ≥ 500 ml threshold.

**Methods:** A decision tree model was developed to assess the cost per disability-adjusted life years (DALYs) averted from a healthcare provider perspective when the World Health Organization (WHO) first-response treatment bundle was initiated using alternative criteria. Sensitivity and specificity of scenarios for identifying women at high risk of a composite outcome of maternal mortality or severe morbidity was estimated using a WHO individual participant data meta-analysis. Direct medical costs (2022 US$) were derived from randomised trial data in Kenya, Nigeria, South Africa, and Tanzania.

**Findings:** Treatment initiation based on lower blood loss thresholds could avert additional composite outcomes. Use of lower blood loss thresholds in combination with any other abnormal haemodynamic marker was more cost-effective than blood loss alone. Combining blood loss thresholds (stepwise from 450 to 300 ml) with any other abnormal haemodynamic marker could avert 15–27% of composite outcomes, increase costs by 9–30% per woman, and have a cost of US$260–479 per DALY averted, compared to using 500 ml blood loss alone. Scenarios were more cost-effective for vaginal births, and cost-saving for populations with a composite outcome incidence ≥ 9%.

**Interpretation:** Expanding the conventional criteria for initiating PPH treatment to include lower blood loss thresholds in combination with any other abnormal haemodynamic marker is cost-effective in improving maternal health outcomes, particularly for obstetric populations with high incidence of PPH mortality and severe morbidity. Further research is warranted in women undergoing caesarean birth.

**Funding:** This study was funded by The Gates Foundation (INV-063940) and the UNDP/UNFPA/UNICEF/WHO/World Bank Special Programme of Research, Development and Research Training in Human Reproduction (HRP), a co-sponsored programme executed by the World Health Organization.

**Panel: Research in context:** *Evidence before this study:* A threshold of 500 ml or more blood loss after childbirth has conventionally been used to initiate treatment for postpartum haemorrhage (PPH). Lower blood loss thresholds in combination with any other abnormal haemodynamic marker have been proposed as newer alternative diagnostic criteria to prompt earlier treatment and potentially improve maternal outcomes. However, such approaches risk overtreatment and higher costs which are critical considerations in low-resource settings where the burden of PPH morbidity and mortality is greatest. Until recently, limited evidence on the prognostic accuracy of clinical markers of postpartum bleeding in predicting maternal mortality and morbidity prevented robust cost-effectiveness analyses of diagnostic criteria for PPH treatment initiation. In 2025, the World Health Organization (WHO) concluded a prognostic accuracy study based on individual participant data (IPD) meta-analysis including 312 151 women from 12 datasets. The prognostic accuracy study estimated the sensitivity and specificity of clinical markers of postpartum bleeding (blood loss, pulse, systolic and diastolic blood pressure, and shock index) in predicting a composite outcome of maternal mortality or severe morbidity (blood transfusion, surgical intervention to stop bleeding, or intensive care admission), providing the first opportunity to evaluate the economic implications of alternative criteria for initiating PPH treatment compared to the conventional 500 ml or more blood loss threshold.

*Added value of this study:* Building on the WHO IPD meta-analysis, we assessed the cost-effectiveness of initiating PPH treatment at lower blood loss thresholds (stepwise from 450 ml, 400 ml, 350 ml, 300 ml), alone and in combination with any other abnormal haemodynamic marker (defined as abnormal pulse rate, systolic or diastolic blood pressure, or shock index), compared with the conventional ≥ 500 ml blood loss threshold. Where treatment initiation was based on lower blood loss thresholds and any abnormal haemodynamic marker, the 500 ml blood loss threshold was retained as an additional criterion, to ensure that women who continue to bleed in the absence of any other abnormal haemodynamic marker still receive treatment as per current convention. We found that lower blood loss thresholds could avert additional composite outcomes, but diagnostic criteria combining lower blood loss thresholds with any other abnormal haemodynamic marker were more cost-effective than those based on blood loss alone, owing to higher specificity and reduced overtreatment. Compared with 500 ml blood loss alone, the cost per disability-adjusted life year (DALY) averted for treatment initiation at 300 ml plus any other abnormal haemodynamic marker (US$479) represents less than 100%, 50%, and 25% of gross domestic product (GDP) per capita for 97%, 85%, and 70% of low- and middle-income countries. For vaginal births, the cost per DALY averted (US$77) was less than 25% of GDP per capita for 99% of low- and middle-income countries. In high-risk obstetric populations with ≥ 9% incidence of the composite outcome, all scenarios combining blood loss with any other abnormal haemodynamic marker were cost-saving (compared to 2.5% incidence in the IPD meta-analysis).

*Implications of all the available evidence:* From a health economics perspective, our findings support expanding diagnostic criteria for initiating PPH treatment to include blood loss thresholds lower than 500 ml in combination with any other abnormal haemodynamic marker. This strategy improves maternal outcomes at acceptable or favourable cost-effectiveness ratios, particularly in low-resource settings with high burden of life-threatening PPH complications. As most women in the WHO IPD meta-analysis had vaginal births, further research is warranted to evaluate the cost-effectiveness of expanded treatment initiation criteria for caesarean births.

## Introduction

Excessive bleeding after birth, known as postpartum haemorrhage (PPH), is a leading cause of maternal death worldwide.^1^ The World Health Organization (WHO) first-response PPH treatment bundle comprising uterine massage, administration of uterotonics, tranexamic acid, intravenous fluids, and systematic examination of genital tract with escalation of care as required has been shown to reduce PPH-related morbidity and mortality^2^ and is cost-effective.^3^ However, delays in diagnosis and treatment, together with limited access to medicines, supplies, and trained health workers, continue to contribute to preventable deaths, particularly in low-resource settings.^4^

Conventionally, a threshold of 500 ml or more of blood loss within 24 hours of birth has been used to initiate PPH treatment based on the widely accepted definition.^5,6^ Lower thresholds have recently been proposed to enable earlier intervention and improve maternal outcomes.^7^ However, expanding diagnostic criteria for initiating PPH treatment to include lower blood loss thresholds carries risks of overtreatment and increased health care costs – two key considerations for low resource settings where the burden of PPH-related morbidity and mortality is also highest. These trade-offs can be framed as a health economics question and assessed using cost-effectiveness analysis.

In 2025, the WHO concluded the conduct of an individual participant data (IPD) meta-analysis to assess the prognostic accuracy of clinical markers of postpartum bleeding in predicting the composite outcome of maternal mortality or severe morbidity including blood transfusion, surgical interventions to stop bleeding, or admission to intensive care unit.^8^ We used the prognostic accuracy estimates from this IPD meta-analysis to assess the cost-effectiveness of initiating PPH treatment at lower blood loss thresholds (incrementally decreasing from 450 ml to 300 ml), alone and in combination with any other abnormal haemodynamic marker (abnormal pulse rate, systolic and diastolic blood pressure, or shock index), compared to initiating treatment at the conventional threshold of 500 ml blood loss alone.

## Methods

### Study population

The study population for this cost-effectiveness analysis was the same as the WHO IPD meta-analysis with details published elsewhere.^8^ Briefly, the IPD meta-analysis included 12 datasets comprising 312 151 women. Most women gave birth vaginally (98.1%). Three randomised controlled trials accounted for more than 80% of included women. The largest trial was the E-MOTIVE cluster randomised trial which evaluated the impact of early detection and a first-response treatment bundle, and included 210 132 women from Kenya, Nigeria, South Africa and Tanzania.^2^ The second largest was the CHAMPION trial^10^ which randomized 29 645 women from Argentina, Egypt, India, Kenya, Nigeria,

Singapore, South Africa, Thailand, Uganda and the United Kingdom to either heat-stable carbetocin or oxytocin for prevention of PPH. The third largest trial examined active management of the third stage of labour with or without controlled cord traction in 24 390 women from Argentina, Egypt, India, Kenya, the Philippines, South Africa, Thailand and Uganda.^11^ The other nine studies included five randomized trials, two prospective cohorts, a before and after implementation study and a nested observational study within a randomised trial, with each contributing between 270 and 18 530 participants who were collectively from both low- and high income countries (Argentina, Brazil, China, Egypt, France, Ireland, Kenya, Nigeria, Pakistan, South Africa, Switzerland, Tanzania, Thailand, Vietnam).

### Scenarios considered for PPH treatment initiation

This cost-effectiveness analysis considered criteria for PPH treatment initiation defined by decreasing blood loss thresholds (500 ml, 450 ml, 400 ml, 350 ml, 300 ml) either with or without any other abnormal haemodynamic marker (one or more of systolic blood pressure < 100 mmHg, diastolic blood pressure <60 mmHg, pulse rate > 100 beats per minute and shock index > 1.0) (*Table 1*).

**Table 1:** Diagnostic criteria for postpartum haemorrhage treatment initiation considered in this cost-effectiveness analysis, and their prognostic sensitivity and specificity for identifying women at risk of the composite outcome (source^8^).

| Scenario name | Diagnostic criteria for treatment initiation | Prognostic sensitivity for the composite outcome | Prognostic specificity for the composite outcome |
| --- | --- | --- | --- |
| ≥ 500 ml blood loss (comparator) | Blood loss ≥ 500 ml | 75.7 (60.3, 86.4) | 81.4 (70.7, 88.8) |
| ≥ 450 ml blood loss | Blood loss ≥ 450 ml | 76.5 (63.1, 86.1) | 78.0 (66.4, 86.4) |
| ≥ 400 ml blood loss | Blood loss ≥ 400 ml | 78.9 (66.5, 87.5) | 71.2 (57.0, 82.2) |
| ≥ 350 ml blood loss | Blood loss ≥ 350 ml | 79.8 (68.6, 87.7) | 65.9 (51.0, 78.2) |
| ≥ 300 ml blood loss | Blood loss ≥ 300 ml | 83.9 (72.8, 91.1) | 54.8 (38.0, 70.5) |
| ≥ 450 ml blood loss + clinical marker(s) | 450-499 ml blood loss + any other abnormal haemodynamic marker; or 500 ml alone* | 86.9 (74.2, 93.9) | 76.1 (56.7, 88.5) |
| ≥ 400 ml blood loss + clinical marker(s) | 400-499 ml blood loss + any other abnormal haemodynamic marker; or 500 ml alone* | 87.4 (75.1, 94.1) | 73.1 (52.1, 87.1) |
| ≥ 350 ml blood loss + clinical marker(s) | 350-499 ml blood loss + any other abnormal haemodynamic marker; or 500 ml alone* | 87.4 (75.4, 94.0) | 70.6 (48.5, 86.0) |
| ≥ 300 ml blood loss + clinical marker(s) | 300-499 ml blood loss + any other abnormal haemodynamic marker; or 500 ml alone* | 87.9 (76.1, 94.3) | 66.6 (43.5, 83.7) |

\*Point system (≥ 2 points) (blood loss ranging from lower bound to 499 ml = 1 point; pulse > 100 bpm or systolic < 100 mmHg or diastolic blood pressure < 60 mmHg or Shock Index > 1.0 = 1 point; blood loss ≥ 500 ml = 2 points)

Where treatment initiation was based on both a blood loss threshold and the presence of one or more abnormal haemodynamic marker, ≥ 500 ml blood loss alone was retained as an additional criterion, to ensure that women who continue to lose blood in the absence of any abnormal haemodynamic marker still receive appropriate treatment as per current convention. Hence all scenarios assessed were expansions of the conventional treatment threshold of 500 ml or more blood loss.

Scenarios assessed were developed through a WHO technical consultation involving 26 experts from all WHO regions, and the sensitivity and specificity of each modelled scenario for predicting women at high risk of the composite outcome was taken directly from the IPD meta-analysis (*Table 1*).^8^

### Cost-effectiveness model

We implemented a decision tree model in TreeAge Pro 2024^9^ (Figure 1). For each set of diagnostic criteria for treatment initiation, women in the model were classified as true positive, true negative, false positive, or false negative depending on the sensitivity and specificity of the criteria being used. Women who were diagnosed positive were assumed to receive treatment, which was either unnecessary (false positive) or necessary (true positive). Where treatment was necessary, it was modelled to be successful or unsuccessful in preventing a composite outcome based on estimated treatment effectiveness parameters from the literature (Table *2*). Women who did not receive treatment either experienced the composite outcome (false negative) or did not (true negative).

**Figure 1:**
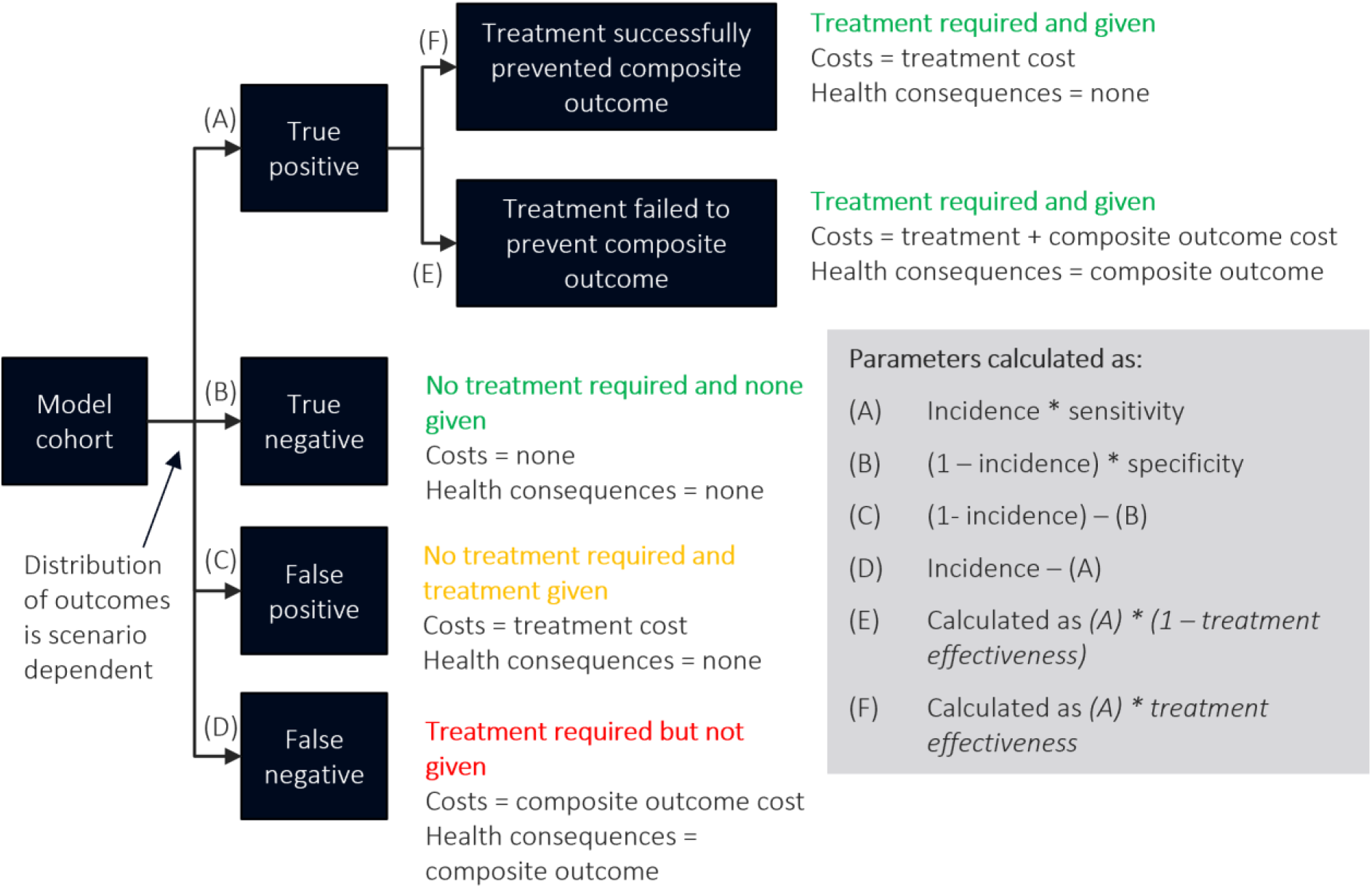
Model schematic. Each set of diagnostic criteria for postpartum haemorrhage treatment initiation has a different prognostic sensitivity and specificity for the composite outcome (defined as one or more of blood transfusion, surgery of any type, morbidity or maternal death). Treatment effectiveness for preventing the composite outcome is estimated from the literature (see Table 2).

**Table 2:** Model parameters.

| Parameter | Value | Source |
| --- | --- | --- |
| Modelled treatment effectiveness (risk ratio for composite outcome vs no treatment)<br>300 ml<br>350 ml<br>400 ml<br>450 ml<br>500 ml | 0.33 (0.20, 0.73)<br>0.36 (0.21, 0.74)<br>0.38 (0.24, 0.78)<br>0.40 (0.26, 0.80)<br>0.43 (0.32, 0.61) | Based on three components:<br>The impact of oxytocin vs placebo or no treatment: risk ratio 0.60 (95%CI 0.41-0.87) for blood transfusion<sup>12</sup><br>The E-MOTIVE bundle of interventions vs usual care: risk ratio 0.71 (95%CI 0.55-0.90) for postpartum blood transfusion<sup>2</sup><br>The impact of earlier treatment on composite outcome: RR = 0.78 (95%CI 0.38-1.59) (see network analysis for 300 ml vs 500 ml,<sup>13</sup> linearly extrapolated to other scenarios).<br>The uncertainty range in value column is the outcome of the multivariate probabilistic sampling during model simulations, with the three effect estimates above parametrized using triangle distributions. |
| Mean cost per woman experiencing the priority composite outcome (2022 US\$)* | \$74.87 | Source: E-MOTIVE cost-effectiveness analysis<sup>3</sup><br>Numerator: From supplementary Table 2, the number of patients in the E-MOTIVE arm (N=48,678) multiplied by the sum of the mean per-patient costs for: laparotomy (mean=\$0.003, S.D=\$0.672), hysterectomy (mean=\$0.117, S.D=\$10.875), non-pneumatic anti shock garment (mean=\$0.002, S.D=\$0.051), bimanual compression (mean=\$0.003, S.D=\$0.093), uterine balloon tamponade (mean=0.002, S.D=\$0.076), blood transfusion (mean=\$1.348, S.D=12.710), ICU admission (mean=\$0.037, S.D=\$3.996), transfer to higher level facility (mean=\$0.089, S.D=\$2.2017), severe PPH (doctor time) (mean=\$0.034, S.D=\$0.334).<br>Denominator: women who had blood transfusion, as proxy for priority composite outcome (N=1063).<br>Uncertainty: in multivariate probabilistic sampling, individual cost components in the numerator were parametrised as lognormal distributions based on the mean and standard deviation for patient costs reported in Supplementary Table 2.<br>Note: Costs are means across E-MOTIVE trial participants, who are from Kenya, Nigeria, South Africa and Tanzania. |
| Mean cost for treatment (excluding composite outcome) (2022 US\$)* | \$6.78 | Source: E-MOTIVE cost-effectiveness analysis<sup>3</sup><br>Numerator: From supplementary Table 2, the number of patients in the E-MOTIVE arm (N=48 678) multiplied by the sum of the mean per-patient costs for: oxytocin (mean=\$0.151, S.D.=\$0.408), tranexamic acid (mean=\$0.367, S.D.=\$1.000), intravenous fluids (mean=\$0.216, S.D.=\$0.582), misoprostol (mean=\$0.066, S.D.=\$0.280), ergometrine (mean=\$0.002, S.D.=\$0.037).<br>Denominator: women who had a uterine massage as a proxy for treatment (N=5762)<br>Uncertainty: in multivariate probabilistic sampling, individual cost components in the numerator were parametrised as lognormal distributions based on the mean and standard deviation for patient costs reported in Supplementary Table 2.<br>Note: Costs are means across E-MOTIVE trial participants, who are from Kenya, Nigeria, South Africa and Tanzania. |
| Components of disability-adjusted life year calculations |  |  |
| *Disutility weight: surgery, transfer to higher level care, or admission to intensive care units* | 0.3242 (0.2197–0.4418) | This is the disutility weight for “PPH > 1000 ml” from the Institute of Health Metrics and Evaluation,<sup>14</sup> assuming that these outcomes are at least as severe as > 1000 ml blood loss. Assumed uniform distribution when sampling. |
| *Disutility weight: blood transfusion* | 0.1488 (0.1007–0.2091) | Institute of Health Metrics and Evaluation<sup>14</sup> disutility weight for severe anaemia due to maternal haemorrhage. Assumed uniform distribution when sampling. |
| *Disutility weight: maternal sepsis* | 0.1334 (0.0885–0.1895) | Institute of Health Metrics and Evaluation.<sup>14</sup> Assumed uniform distribution when sampling. |
| *Duration PPH disutility applied for* | 6 weeks | Consistent with E-MOTIVE cost-effectiveness analysis<sup>3</sup> |
| *Median age of maternal death (and inter-quartile range)* | 29.0 (24.0–34.0) years | From the harmonized dataset<sup>8</sup> |
| *Among women with composite outcome, maternal deaths* | 1.62% | From the harmonized dataset.<sup>8</sup> Among 311 994 with available variables for composite outcomes, there were 7 769 composite outcomes and 126 maternal deaths. |
| *Female health-adjusted years of life remaining at age of PPH death* | 38 (34–42) years | Female health-adjusted years of life remaining by country at age of maternal death (29 [24–34] years) taken from Institute of Health Metrics and Evaluation 2021 Global Burden of Disease analysis.<sup>15</sup> Weighted average across countries according to estimates of total PPH deaths in each country (in 2023).<sup>16</sup> |
| *Discounted years of life lost per maternal death* | 23.17 (21.77–24.41) | 38 (34–42) years with 3% per annum applied to future years. |
| *Mean disability-adjusted life years per woman experiencing the composite outcome* | 0.3949 (0.3257–0.4683) | Estimated as disutility weight from the composite outcome plus years of life lost from maternal mortality. Uncertainty intervals were estimated using bootstrap for uncertainty in number of maternal deaths. Each bootstrap iteration disutility weights and age of death were sampled from uniform distributions from their ranges stated above. |
| Incidence of composite outcome in absence of treatment | 25 per 1000 women | From the harmonised dataset.<sup>8</sup> Among 311 994 with available variables for composite outcome, there were 7 769 composite outcomes. |
| Cohort size used for microsimulations |  | Number of women in harmonised dataset with sufficiently complete data for analysis / sub-analysis.<sup>8</sup> |
| *Blood loss models* | 305 523 |  |
| *Multivariate models* | 30 191 |  |
| *Blood loss models (vaginal births only)* | 300 214 |  |
| *Multivariate models (vaginal births only)* | 26 682 |  |

\* Costs relate to low resource settings, but two-way sensitivity analyses were conducted for a wide range of unit costs for treatment and for the composite outcome.

### Treatment effectiveness

The model considered treatment with the WHO first-response treatment bundle.^2^ Direct effectiveness estimates that compare this treatment bundle to no treatment are not available. Therefore, as advised by the WHO technical experts consultation, treatment effectiveness compared to no treatment was estimated by multiplying the impact of oxytocin versus placebo or no treatment (risk ratio 0.60 [95%CI 0.41-0.87] for blood transfusion^12^) and the E-MOTIVE bundle of interventions versus usual care (risk ratio 0.71 [95%CI 0.55-0.90] for postpartum blood transfusion^2^). Effect sizes for reducing blood transfusion were then used as a proxy for treatment effects on the composite outcome, since in 93.0% of cases in the IPD the composite outcome was observed due to blood transfusion only).^8^

In addition to more women receiving treatment in some scenarios, we assumed the treatment can be more effective if initiated earlier. This additional impact of earlier treatment was included for blood loss thresholds less than 500 ml, based on the effect of 300 ml versus 500 ml diagnosis threshold on the composite outcome from a network analysis (RR = 0.78 [95%CI 0.38-1.59]^13^), linearly extrapolated to the blood loss thresholds between 300 ml and 500 ml. A sensitivity analysis was run excluding this effect. Resulting treatment effectiveness by blood loss diagnostic criteria are in Table 2.

### Cost data

The unit cost of treatment (excluding the composite outcome), and the mean cost per woman experiencing the composite outcome were estimated from a healthcare provider’s perspective, based on the E-MOTIVE trial cost-effectiveness analysis^3^ (Table 2). These reflect costs across Kenya, Nigeria, South Africa and Tanzania where the study was conducted. All costs are presented in 2022 US$ and without discounting due to the short analysis time frame.

### Disability-adjusted life years per composite outcome

Mean DALYs per woman experiencing the composite outcome were estimated from years of life lost from maternal death and years lived with disability from sub-components of the composite outcome (Table 2). Years of life lost were based on median age of death from the IPD and health-adjusted years of life remaining at age of death,^15^ weighted across countries based on the global distribution of PPH deaths,^16^ with 3% per annum discounting applied to future years. Where disutility weights were not available for sub-components that were procedures (as opposed to health outcomes), they were approximated based on morbidities commensurate with those procedures. For women with any surgical intervention to stop bleeding or admission to intensive care units, the disutility weight for “PPH > 1000 ml” was used (assuming that these outcomes are at least as severe as PPH > 1000 ml).^14^ For women with blood transfusions, the disutility weight for severe anaemia due to maternal haemorrhage was used. For women with sepsis there was a corresponding disutility weight. Disutility weights were applied for a six-week duration.^3^ Women who experienced multiple sub-components were assigned the maximum of the corresponding disutility weights. Bootstrap was used to generate 95% uncertainty intervals (95%UIs) for mean DALYs per woman experiencing the composite outcome. For each bootstrap iteration, disutility weights were re-sampled from their reported uncertainty range.^14^

### Outcomes and analysis

For each treatment initiation criteria, the model projected the number of composite outcomes per 1000 women, number of unnecessary treatments (i.e. false positives) per 1000 women, mean cost per woman, and mean DALYs lost per woman. The incremental cost-effectiveness ratio (ICER; cost per DALY averted) was calculated for each scenario compared to the 500 ml blood loss only scenario.

### Subgroup analysis

A subgroup analysis was conducted considering vaginal births only.

### Uncertainty analyses

Multivariate probabilistic uncertainty analyses were conducted for each scenario to estimate 95%UIs. Parameters for sensitivity, specificity and treatment effectiveness were sampled 1000 times (using triangle distributions), and for each parameter set microsimulations were run to estimate patient outcomes. The number of microsimulations per parameter set was equal to the number of women in the IPD with sufficiently complete data to inform the sensitivity and specificity estimates (Table 2). Unit costs and DALYs were sampled at an individual level within each microsimulation (using lognormal distributions for component costs and a uniform distribution for DALYs). Uncertainty distributions for parameters are described in Table 1 and Table 2.

For different thresholds of willingness-to-pay per DALY averted, the preferred strategy was estimated as the treatment initiation criteria that maximised net benefits^17^ (i.e. *DALYs averted * willingness-to-pay – total costs*) for the greatest proportion of the 1000 model runs.

### Sensitivity analyses

Sensitivity analyses were conducted considering populations with incidence of the composite outcome ranging from 2% to 10% (e.g. in some settings with high prevalence of anaemia incidence of up to 25% of postpartum blood transfusion has been observed^18^), rather than the 2.5% in the IPD, and excluding the impact of early treatment (Appendix A). Two-way sensitivity analyses were undertaken considering point estimate outcomes for wide ranges of treatment and composite outcome unit costs, since these are likely to vary substantively across countries (Appendix B).

### Ethics

Eligible studies for the WHO IPD meta-analysis^8^ must have had prior ethics committee approval and the data holder had to be able to enter into a legal data sharing agreement with WHO. Institutional review board approval was not required for this study because only previously collected, de-identified data were used. All contributing studies had received appropriate ethical approvals, and no new participant recruitment or data collection was undertaken for the current study.

## Results

Using 500 ml blood loss only (current standard-of-care), with treatment applied to true positives there were 14 (95%UI 11–18) composite outcomes and 5.6 (95%UI 4.5–7.1) DALYs lost per 1000 women, with a mean cost per woman of $2.40 (95%UI 2.02–3.06) (Figure 2 and Table 3). The incidence of composite outcomes reduced in all scenarios compared to the 500 ml blood loss scenario (Figure 2, top left). However, the costs were also higher for each scenario (Figure 2, top right), meaning that the additional treatment costs were not completely offset by costs saved from additional composite outcomes averted.

**Figure 2:**
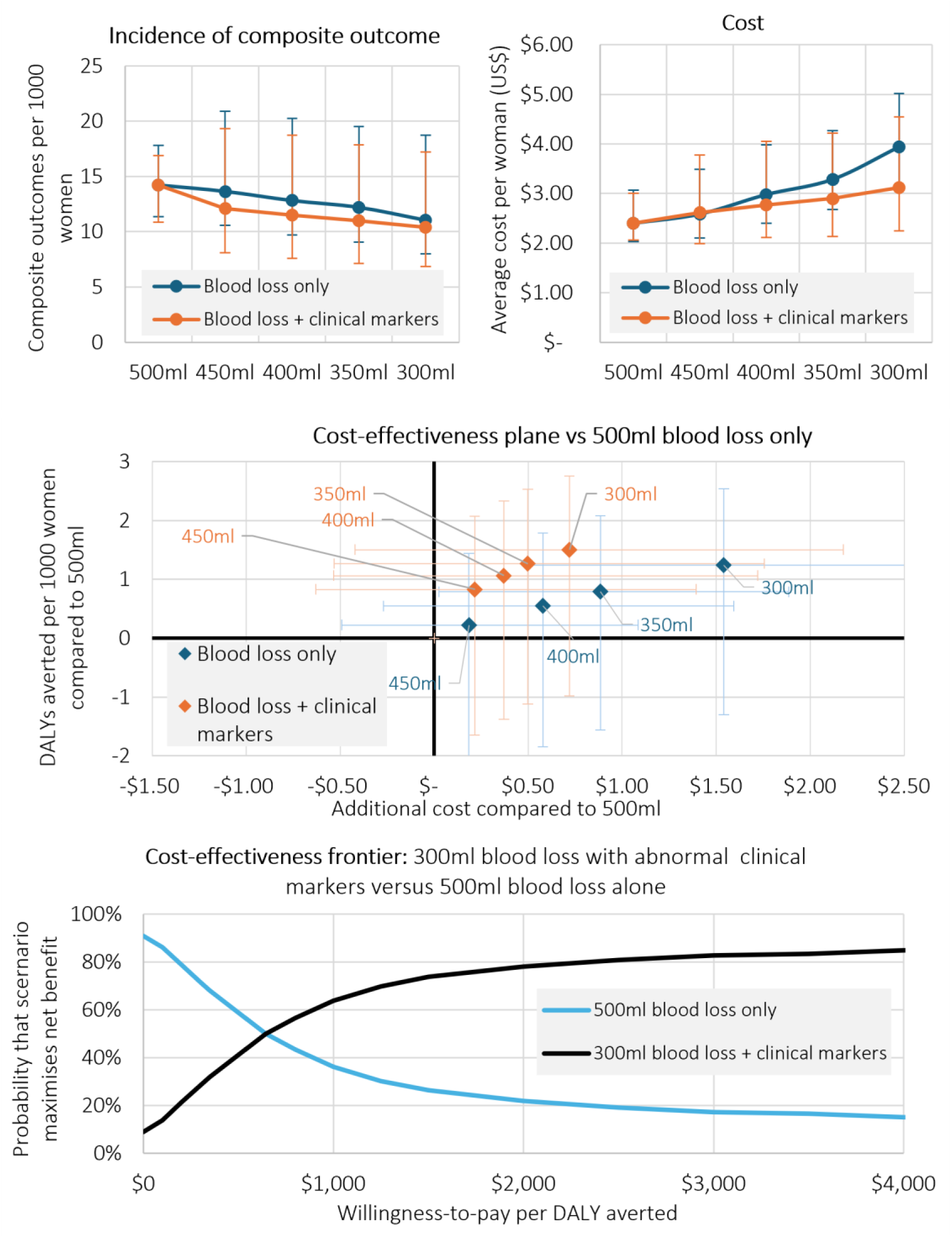
Model outcomes for blood loss thresholds alone (“blood loss only”), or blood loss thresholds in combination with any other abnormal haemodynamic marker (“blood loss + clinical markers”). Top left: incidence of composite outcome per 1000 women; top right: mean cost per woman (in 2022 US$); middle: disability-adjusted life years (DALYs) averted per 1000 women and difference in mean cost for scenarios compared to the 500 ml blood loss threshold; bottom: cost-effectiveness frontier, showing the probability that a scenario maximises net benefits for a given willingness-to-pay per DALY averted.

**Table 3:** Modelled impact of scenarios on incidence of the composite outcome, disability-adjusted life years (DALYs) and mean costs, as well as the difference in outcomes compared to the 500 ml blood loss threshold.

| Scenario | Incidence of composite outcome per 1000 women | Mean DALYs lost per 1000 women | Mean cost per woman (2022 US\$) | Unnecessary treatments per 1000 women | Difference in composite outcomes per 1000 women, compared to 500 ml blood loss | Difference in DALYs per 1000 women, compared to 500 ml blood loss | Mean additional cost per woman compared to 500 ml blood loss (2022 US\$) | Cost per DALY averted compared to 500 ml blood loss (2022 US\$) |
| --- | --- | --- | --- | --- | --- | --- | --- | --- |
| **Blood loss** |  |  |  |  |  |  |  |  |
| ≥ 500 ml blood loss | 14.2<br>(11.4, 17.8) | 5.6<br>(4.5, 7.1) | \$2.40<br>(\$2.02, \$3.06) | 189<br>(127, 261) | | | | |
| ≥ 450 ml blood loss | 13.6<br>(10.6, 20.9) | 5.4<br>(4.2, 8.3) | \$2.58<br>(\$2.10, \$3.49) | 222<br>(152, 304) | -0.6<br>(-3.6, 5.3) | -0.2<br>(-1.4, 2.1) | \$0.18<br>(-\$0.49, \$1.09) | \$826<br>(-\$6193, \$4593) |
| ≥ 400 ml blood loss | 12.8<br>(9.7, 20.2) | 5.1<br>(3.8, 8.0) | \$2.98<br>(\$2.40, \$3.98) | 289<br>(196, 396) | -1.4<br>(-4.5, 4.7) | -0.5<br>(-1.8, 1.8) | \$0.58<br>(-\$0.27, \$1.59) | \$1055<br>(-\$8514, \$12054) |
| ≥ 350 ml blood loss | 12.2<br>(9.0, 19.5) | 4.8<br>(3.6, 7.8) | \$3.28<br>(\$2.67, \$4.27) | 342<br>(235, 448) | -2.0<br>(-5.2, 3.9) | -0.8<br>(-2.1, 1.6) | \$0.88<br>(\$0.03, \$1.89) | \$1122<br>(-\$14607, \$11770) |
| ≥ 300 ml blood loss | 11.0<br>(8.0, 18.7) | 4.4<br>(3.2, 7.4) | \$3.94<br>(\$3.11, \$5.02) | 444<br>(324, 574) | -3.1<br>(-6.4, 3.3) | -1.2<br>(-2.5, 1.3) | \$1.54<br>(\$0.50, \$2.59) | \$1240<br>(-\$12986, \$15133) |
| **Blood loss + clinical marker(s)** |  |  |  |  |  |  |  |  |
| ≥ 450 ml blood loss + clinical marker(s) | 12.1<br>(8.1, 19.3) | 4.8<br>(3.3, 7.6) | \$2.61<br>(\$1.99, \$3.77) | 253<br>(142, 387) | -2.1<br>(-5.3, 4.1) | -0.8<br>(-2.1, 1.6) | \$0.22<br>(-\$0.63, \$1.39) | \$260<br>(-\$6458, \$8853) |
| ≥ 400 ml blood loss + clinical marker(s) | 11.5<br>(7.6, 18.7) | 4.5<br>(3.1, 7.4) | \$2.77<br>(\$2.11, \$4.05) | 287<br>(160, 433) | -2.7<br>(-5.9, 3.5) | -1.1<br>(-2.3, 1.4) | \$0.37<br>(-\$0.53, \$1.72) | \$349<br>(-\$7921, \$7727) |
| ≥ 350 ml blood loss + clinical marker(s) | 11.0<br>(7.1, 17.9) | 4.3<br>(2.9, 7.1) | \$2.90<br>(\$2.14, \$4.22) | 298<br>(169, 459) | -3.2<br>(-6.5, 2.8) | -1.3<br>(-2.5, 1.1) | \$0.50<br>(-\$0.53, \$1.76) | \$393<br>(-\$8213, \$7367) |
| ≥ 300 ml blood loss + clinical marker(s) | 10.4<br>(6.8, 17.2) | 4.1<br>(2.8, 6.8) | \$3.12<br>(\$2.24, \$4.54) | 345<br>(198, 506) | -3.8<br>(-7.0, 2.5) | -1.5<br>(-2.8, 1.0) | \$0.72<br>(-\$0.42, \$2.18) | \$479<br>(-\$9325, \$7758) |

The diagnostic criteria for treatment initiation that combined lower blood loss thresholds with any other abnormal haemodynamic marker had a lower cost per DALY averted than those based on blood loss only, primarily due to their higher specificity leading to fewer unnecessary treatments. For example, compared to initiating treatment using a threshold of 500 ml blood loss alone, using a threshold of 300 ml blood loss alone cost an additional $1.54 (95%UI 0.50–2.59) per woman (+64%) and averted 3 (95%UI −3–6) additional composite outcomes per 1000 women, at a cost of $1,240 (95%UI −12,986–15,133) per DALY averted. In contrast, using a threshold of 300 ml in combination with any other abnormal haemodynamic marker cost an additional $0.72 (95%UI −0.42–2.18) per woman (+30%) and averted 4 (95%UI −3–7) additional composite outcomes per 1000 women, at a cost of $479 (95%UI −9,325–7,758) per DALY averted compared to initiating treatment using a threshold of 500 ml blood loss alone (Table 3). An ICER of $479 represents between 0.03- and 1.91- times GDP per capita across low- and middle-income countries (Supplementary Table C1).

There was substantial uncertainty associated with outcomes, owing to the wide confidence intervals of model parameters (Figure 2, middle). When the threshold of 300 ml blood loss in combination with any other abnormal haemodynamic marker was compared to the threshold of 500 ml blood loss alone, it was the preferred strategy at a willingness-to-pay of >$645 per DALY averted (Figure 2, bottom). Comparisons against the full set of scenarios are in Supplementary Figure C1.

The expanded diagnostic criteria for treatment initiation had higher specificity and were more cost-effective for vaginal births than for all births (Table 4, Supplementary Table A1). For example, for vaginal births only, the threshold of 300 ml blood loss in combination with any other abnormal haemodynamic marker had a cost of only $77 (95%UI −3,920–5,385) per DALY averted compared to 500 ml blood loss alone. An ICER of $77 represents between 0.005- and 0.31-times GDP per capita across low- and middle-income countries (Supplementary Table C1). For vaginal births, when the threshold of 300 ml blood loss in combination with any other abnormal haemodynamic marker was compared to the threshold of 500 ml blood loss alone, it was the preferred strategy for willingness-to-pay of > $230 per DALY averted (Supplementary Figure A1).

**Table 4:** Results for vaginal birth sub analysis. Modelled impact of scenarios on incidence of the composite outcome, disability-adjusted life years (DALYs) and mean costs, as well as the difference in outcomes compared to the 500 ml blood loss threshold.

| Scenario | Incidence of composite outcome per 1000 women | Mean DALYs lost per 1000 women | Mean cost per woman (2022 US\$) | Unnecessary treatments per 1000 women | Difference in composite outcomes per 1000 women, compared to 500 ml blood loss | Difference in DALYs per 1000 women, compared to 500 ml blood loss | Mean additional cost per woman compared to 500 ml blood loss (2022 US\$) | Cost per DALY averted compared to 500 ml blood loss (2022 US\$) |
| --- | --- | --- | --- | --- | --- | --- | --- | --- |
| **Blood loss** |  |  |  |  |  |  |  |  |
| ≥ 500 ml blood loss | 14.6<br>(11.8, 18.2) | 5.8<br>(4.7, 7.2) | \$2.05<br>(\$1.81, \$2.50) | 129<br>(99, 163) | | | | |
| ≥ 450 ml blood loss | 14.1<br>(10.9, 21.0) | 5.6<br>(4.3, 8.4) | \$2.17<br>(\$1.87, \$2.86) | 153<br>(120, 189) | -0.5<br>(-3.6, 5.1) | -0.2<br>(-1.4, 2.0) | \$0.12<br>(-\$0.27, \$0.73) | \$593<br>(-\$3235, \$3528) |
| ≥ 400 ml blood loss | 13.3<br>(10.2, 20.4) | 5.2<br>(4.1, 8.1) | \$2.45<br>(\$2.07, \$3.20) | 203<br>(153, 260) | -1.4<br>(-4.5, 4.6) | -0.5<br>(-1.8, 1.8) | \$0.40<br>(-\$0.10, \$1.08) | \$740<br>(-\$9042, \$5984) |
| ≥ 350 ml blood loss | 12.6<br>(9.5, 19.6) | 5.0<br>(3.8, 7.8) | \$2.68<br>(\$2.28, \$3.43) | 246<br>(186, 304) | -2.0<br>(-5.2, 3.8) | -0.8<br>(-2.1, 1.5) | \$0.63<br>(\$0.12, \$1.31) | \$805<br>(-\$10390, \$10146) |
| ≥ 300 ml blood loss | 11.7<br>(8.6, 19.0) | 4.6<br>(3.4, 7.5) | \$3.28<br>(\$2.67, \$4.23) | 344<br>(254, 446) | -2.9<br>(-6.1, 3.3) | -1.2<br>(-2.4, 1.3) | \$1.23<br>(\$0.51, \$2.08) | \$1065<br>(-\$12772, \$13598) |
| **Blood loss + clinical marker(s)** |  |  |  |  |  |  |  |  |
| ≥ 450 ml blood loss + clinical marker(s) | 12.9<br>(8.9, 19.6) | 5.1<br>(3.6, 7.8) | \$2.04<br>(\$1.82, \$2.57) | 143<br>(113, 172) | -1.7<br>(-5.1, 4.1) | -0.7<br>(-2.0, 1.6) | -\$0.02<br>(-\$0.33, \$0.48) | -\$24<br>(-\$1805, \$2244) |
| ≥ 400 ml blood loss + clinical marker(s) | 12.1<br>(8.3, 19.1) | 4.8<br>(3.3, 7.5) | \$2.08<br>(\$1.86, \$2.64) | 159<br>(125, 191) | -2.5<br>(-6.0, 3.4) | -1.0<br>(-2.4, 1.3) | \$0.03<br>(-\$0.31, \$0.56) | \$29<br>(-\$3764, \$2638) |
| ≥ 350 ml blood loss + clinical marker(s) | 11.3<br>(7.4, 17.8) | 4.5<br>(3.0, 7.0) | \$2.10<br>(\$1.84, \$2.67) | 166<br>(134, 200) | -3.3<br>(-6.6, 2.7) | -1.3<br>(-2.6, 1.1) | \$0.05<br>(-\$0.34, \$0.57) | \$35<br>(-\$3357, \$2708) |
| ≥ 300 ml blood loss + clinical marker(s) | 10.1<br>(6.4, 17.7) | 4.0<br>(2.6, 7.0) | \$2.19<br>(\$1.88, \$2.92) | 195<br>(155, 240) | -4.5<br>(-7.7, 2.9) | -1.8<br>(-3.0, 1.2) | \$0.14<br>(-\$0.28, \$0.82) | \$77<br>(-\$3920, \$5385) |

In populations with ≥ 9% incidence of the composite outcome, diagnostic criteria for treatment initiation that combined lower blood loss thresholds with any other abnormal haemodynamic marker were cost saving compared to 500 ml blood loss alone (Supplementary Figure C3).

With the early treatment effect excluded from the model, the impact of each scenario was reduced, and the cost per DALY averted increased (Supplementary Figure A3, Supplementary Table A3).

The cost per DALY averted increased with higher treatment cost and reduced with higher costs associated with the composite outcome (Supplementary Excel Appendix B).

## Discussion

Our study assessed the cost-effectiveness of different diagnostic criteria for PPH treatment initiation, including use of lower blood loss thresholds with and without consideration of other abnormal haemodynamic markers. Point estimate results suggest that compared to a 500 ml blood loss threshold only, reducing the blood loss threshold to between 450 ml and 300 ml could avert an additional 4-22% of composite outcomes, cost a mean additional 8-64% per woman, and have a cost of $826–1,240 per DALY averted. Reducing the blood loss threshold from 500 ml to between 450 ml and 300 ml and also considering other abnormal clinical markers increased the total impact (an additional 15-27% of composite outcomes averted), reduced additional costs (a mean additional 9-30% per woman) and was more cost-effective with a cost of $260–479 per DALY averted. Scenarios were more cost-effective for vaginal births, and cost saving for high-risk obstetric populations with composite outcome incidence ≥ 9%.

Broadening the diagnostic criteria for PPH treatment initiation by using lower blood loss thresholds led to greater impact, regardless of whether other abnormal haemodynamic markers were considered. This is because treatment was assumed to be beneficial where needed, and the modelling did not account for any potential harms due to unnecessary treatment (i.e., for false positives). Early treatment components such as oxytocic drugs are well tolerated and have low reported side effects relative to the potential catastrophic impact of untreated PPH, and views among women and healthcare providers support a need to do more to prevent and detect PPH.^19^ However, any approach to broaden diagnostic criteria for treatment initiation would increase the scale at which treatment is administered and increase procedures such as examination of genital tract, which can be painful and increase maternal dissatisfaction.^20^ Therefore, monitoring of maternal satisfaction and mental health^20–22^ would be an important consideration for the implementation of a newer PPH diagnostic strategy. Views on the acceptability of overtreatment may also differ depending on the risk of severe morbidity and mortality from PPH (e.g. in low- and middle-income countries versus high-income countries).

Diagnostic criteria that considered blood loss in combination with any other abnormal haemodynamic marker were more cost-effective than those considering blood loss alone, due to reduced unnecessary treatment. The use of other abnormal haemodynamic markers as diagnostic criteria led to higher sensitivity and hence greater impact, as well as lower costs due to higher specificity and fewer unnecessary treatments. A limitation of this analysis is the assumption that the diagnostic criteria were implemented perfectly, whereas in practice the use of multiple criteria adds complexity that may result in imperfect implementation. For example, challenges have been observed previously with the introduction of drapes to objectively measure blood loss.^23^ If new guidelines are adopted, sufficient education and communication will be required and the implications of any increased workloads for healthcare providers will need to be monitored. Follow up should confirm that any changes have been well understood and appropriately implemented by healthcare providers, and that they are associated with an improvement of maternal outcomes.

By definition, a scenario is considered cost-effective if the cost per DALY averted is less than a specified willingness-to-pay threshold. However, there is no consensus on what the willingness-to-pay threshold should be, and it would vary widely across settings. Previous estimates of willingness-to-pay thresholds based on health opportunity costs suggest that thresholds may be less than one GDP per capita for countries with low Human Development Index.^24^ The cost per DALY averted from the 300 ml blood loss in combination with any other abnormal haemodynamic marker was less than 100%, 50% and 25% of GDP per capita for 97% (119/123), 85% (105/123) and 70% (86/123) of low- and middle-income countries, respectively, and for vaginal births was less than 25% of GDP per capita for 99% (122/123) of countries. For higher risk obstetric populations all scenarios that considered other abnormal haemodynamic markers were cost saving. On balance this suggests that the 300 ml blood loss in combination with any other abnormal haemodynamic marker is likely to be cost-effective, because for countries with low Human Development Index where the willingness-to-pay threshold is low relative to per capita GDP, the incidence of the composite outcome is also likely to be higher. For high income countries, treatment costs and composite outcome costs are both likely to be higher, but if the increase in composite outcome costs is greater than the increase in the treatment cost this can potentially enhance the cost-effectiveness of the scenarios modelled.

Assessing which strategies maximise net benefits can inform decision-making by accounting for uncertainty in results. For a given willingness-to-pay threshold, many scenarios may be cost-effective, and the preferred strategy is the one that is both cost-effective and has the greatest total impact for the largest proportion of model simulations. For decision-makers who have a known willingness-to-pay threshold, the preferred strategy can be interpreted as the one that will achieve the greatest impact without exceeding this cost-effectiveness threshold. The main analysis compared just the 300 ml threshold in combination with any other abnormal haemodynamic marker and the 500 ml blood loss scenarios (other scenarios in Supplementary Appendix C), and showed that even accounting for uncertainty, the 300 ml threshold in combination with any other abnormal haemodynamic marker is likely to be the preferrable strategy from a health economics perspective at >$645 per DALY averted for all births, and at >$230 per DALY averted for vaginal births only.

There was a high degree of uncertainty in the estimated impact, cost, and cost-effectiveness of the different scenarios, which is reflected in their wide uncertainty intervals. Practically, there are not likely to be any ways to reduce this uncertainty. On the impact side, the composite outcome is sufficiently rare that the sample size required for higher certainty is impractically large. Data for this analysis was obtained through WHO-led global calls and pooling of available studies, with the resulting sample size (n=312,151) being the largest that could be achieved through this extensive process.^8^ On the cost side there were two major drivers of uncertainty, first through feedback from the uncertain impacts (i.e. how many treatments or composite outcomes there would be to incur costs), and second from the unit cost inputs that had their own large uncertainty intervals,^3^ driven by real-world differences in costs occurred between individuals.

There are some additional limitations to our study. First, there is a bias in the study data where some treatment was initiated among the cohort (i.e. women classified as false positive in the 500 ml comparator scenario may have been true positive but treated effectively). However, this would potentially lead to underestimating benefits, particularly if some women in the dataset had received early effective treatment which was therefore missed from the analysis. Second, cost data was used from the E-MOTIVE trial, which was conducted across Kenya, Nigeria, South Africa and Tanzania. Costs are likely to vary substantively by country and so these results may not be generalizable. Future research should focus on collecting more diverse, localised cost data from a broader range of countries to enhance the generalisability of these findings. Third, costs included in this analysis were limited to treatment and composite outcome costs and did not include the costs of any additional complications. Fourth, a very large majority of women in the datasets had vaginal births, and so further work is warranted for caesarean sections. Finally, the model is a simplification of clinical practice and does not capture the full nuance of identifying and treating PPH. Results should be considered as average outcomes to inform guideline decision-making alongside other evidence domains.

Diagnostic criteria for initiating PPH treatment based on blood loss thresholds in combination with any other abnormal haemodynamic marker are likely to be more cost-effective than using blood loss thresholds alone. When compared to the conventional threshold of 500 ml blood loss alone, the 300 ml blood loss threshold in combination with any other abnormal haemodynamic marker is likely to be cost-effective in a majority of low- and middle-income countries, particularly for vaginal births, and cost saving for populations with very high incidence of the composite outcome. Uncertainty intervals were wide; however, given the large sample size of participant data upon which our study was based, these are unlikely to be practically reduced by further research. Further research is needed evaluate the cost-effectiveness of expanded treatment initiation criteria for caesarean births and for births in high income countries.

## Contributors

IG and OTO conceptualised the study. NS, IG, CRW, AC, OTO designed the methods and analytic approach. NS and TW implemented the model and ran the scenarios. NS, IG, TW, CRW, AC, OTO extracted and validated model parameters. NS wrote the first draft of the manuscript. All authors contributed to analysis and interpretation, and critically revised the manuscript for intellectual content. NS, TW, IG, CRW, and OTO had full access to and verified all the study data. All authors approved the final manuscript and had final responsibility for the decision to submit for publication.

## Data sharing

All model parameters used for this study are available in the manuscript and supplementary material, extracted from the sources as cited. The individual participant data (IPD) that served as the basis for this article were shared with the WHO exclusively for the purpose of conducting this IPD meta-analysis to reappraise the definition of PPH. Researchers interested in accessing data from the original studies should contact the corresponding authors of those studies directly. Access will be subject to the approval of data owners and applicable data sharing agreements.

## Acknowledgements

This study was funded by The Gates Foundation (INV-063940) and the UNDP/UNFPA/UNICEF/WHO/World Bank Special Programme of Research, Development and Research Training in Human Reproduction (HRP), a co-sponsored programme executed by the World Health Organization. The views expressed in this manuscript are those of the named authors and do not necessarily reflect the views of HRP or the World Health Organization. The authors acknowledge the contributions of the Members of the WHO Consortium on Postpartum Haemorrhage Definition (listed in alphabetical order within each group) include: lead investigators and country principal investigators of the original studies that provided included datasets – Anderson Borovac-Pinheiro (University of Campinas, São Paulo, Brazil), Guillermo Carroli (Centro Rosarino de Estudios Perinatales, Rosario, Argentina), Arri Coomarasamy (Tommy’s National Centre for Miscarriage Research, University of Birmingham, UK), Jill Durocher (Gynuity Health Projects, New York, USA), Fadhlun M. Alwy Al-Beity (Muhimbili University of Health and Allied Sciences, Dar es Salaam, Tanzania), Sue Fawcus (University of Cape Town, South Africa), Mario Festin (University of the Philippines, Manila, The Philippines), Hadiza S. Galadanci (Bayero University, Kano, Nigeria), Shivaprasad Goudar (Women’s and Children’s Health Research Unit, Jawaharlal Nehru Medical College, KLE Academy of Higher Education and Research, Belagavi, India), A. Metin Gülmezoglu (Concept Foundation, Geneva, Switzerland), Christian Haslinger (Department of Obstetrics, University Hospital Zurich, University of Zurich, Switzerland), G. Justus Hofmeyr (Effective Care Research Unit, University of the Witwatersrand, Walter Sisulu University, East London, South Africa), Pisake Lumbiganon (Faculty of Medicine, Khon Kaen University, Thailand), Kidza Mugerwa (Makerere University, Kampala, Uganda), Rodolfo C. Pacagnella (University of Campinas, São Paulo, Brazil), Zahida Qureshi (University of Nairobi, Nairobi, Kenya), Loïc Sentilhes (Department of Obstetrics and Gynecology, Bordeaux University Hospital, France), and Lumaan Sheikh (Aga Khan University, Karachi, Pakistan); members of the PPH Definition Evidence Synthesis Team – John Allotey (Institute of Life Course and Medical Sciences, University of Liverpool, Liverpool, UK), Arri Coomarasamy, Adam Devall (Tommy’s National Centre for Miscarriage Research, University of Birmingham, UK), Jonathan J. Deeks (Department of Applied Health Sciences, University of Birmingham, Birmingham, UK), Malcolm Price (Department of Public Health, Canadian University Dubai, Dubai, United Arab Emirates), Soha Sobhy (Tommy’s National Centre for Miscarriage Research, University of Birmingham, UK), Aurelio Tobias (Spanish Council for Scientific Research, Barcelona, Spain) and Idnan Yunas (Department of Metabolism and Systems Science, College of Medicine and Health, University of Birmingham, Birmingham, UK); and the WHO Project Secretariat – Fernando Althabe, Jenny Cresswell, Ioannis Gallos, Olufemi T. Oladapo, and Caitlin R. Williams (United Nations (UN) Development Program–UN Population Fund–UN Children’s Fund–World Health Organization (WHO)– World Bank Special Program of Research, Development, and Research Training in Human Reproduction, Department of Sexual and Reproductive Health and Research, WHO, Geneva, Switzerland). The authors are also grateful for the contributions of participants at the WHO Technical Consultation on the Reappraisal of Postpartum Haemorrhage Definition for Contemporary Clinical Practice, Research and Policy, held on 21-23 January 2025.

