## Supplement for "Cost-effectiveness of diagnostic criteria for postpartum haemorrhage treatment"

### APPENDIX A: sensitivity analyses

**Sub analysis for vaginal births only**

Of the 305 523 individuals in the dataset with complete data, 300 214 were vaginal births. The sensitivity and specificity of the different diagnostic criteria for PPH treatment initiation were estimated to be different for this group, and so a sub analysis was run. The scenario parameters are in Table A1.

As with the main analysis, when only vaginal births were considered, lower blood loss thresholds resulted in greater impact, and scenarios using blood loss in combination with any other abnormal haemodynamic marker were more cost-effective than those using blood loss only. There was still large uncertainty (Figure A1). When the threshold of 300 ml blood loss in combination with any other abnormal haemodynamic marker was compared to the threshold of 500 ml blood loss alone, it was the preferred strategy for willingness-to-pay of > $230 per DALY averted.

Table A1: Vaginal birth sub analysis parameters.

| Scenario name | Diagnostic criteria for treatment initiation | Sensitivity | Specificity |
|---|---|---|---|
| ≥ 500 ml blood loss (comparator) | Blood loss ≥ 500 ml | 72.8 (56.7, 84.6) | 87.0 (82.1, 90.7) |
| ≥ 450 ml blood loss | Blood loss ≥ 450 ml | 73.4 (58.8, 84.3) | 84.6 (79.5, 88.6) |
| ≥ 400 ml blood loss | Blood loss ≥ 400 ml | 76.0 (63.0, 85.6) | 79.6 (72.1, 85.6) |
| ≥ 350 ml blood loss | Blood loss ≥ 350 ml | 77.1 (65.8, 85.5) | 75.4 (67.1, 82.2) |
| ≥ 300 ml blood loss | Blood loss ≥ 300 ml | 80.1 (68.5, 88.2) | 65.3 (51.8, 76.7) |
| ≥ 450 ml blood loss + clinical marker(s) | 450-499 ml blood loss + any other abnormal haemodynamic marker; or 500 ml alone* | 81.2 (64.4, 91.2) | 85.6 (81.3, 89.0) |
| ≥ 400 ml blood loss + clinical marker(s) | 400-499 ml blood loss + any other abnormal haemodynamic marker; or 500 ml alone* | 83.3 (64.3, 93.2) | 84.1 (79.4, 87.9) |
| ≥ 350 ml blood loss + clinical marker(s) | 350-499 ml blood loss + any other abnormal haemodynamic marker; or 500 ml alone* | 85.2 (63.9, 94.9) | 83.0 (78.2, 87.0) |
| ≥ 300 ml blood loss + clinical marker(s) | 300-499 ml blood loss + any other abnormal haemodynamic marker; or 500 ml alone* | 89.9 (57.9, 98.3) | 80.3 (73.9, 85.3) |

*Point system (≥ 2 points) (blood loss ranging from lower bound to 499 ml = 1 point; pulse > 100 bpm or systolic < 100 mmHg or diastolic blood pressure < 60 mmHg or Shock Index > 1.0 = 1 point; blood loss ≥ 500 ml = 2 points)

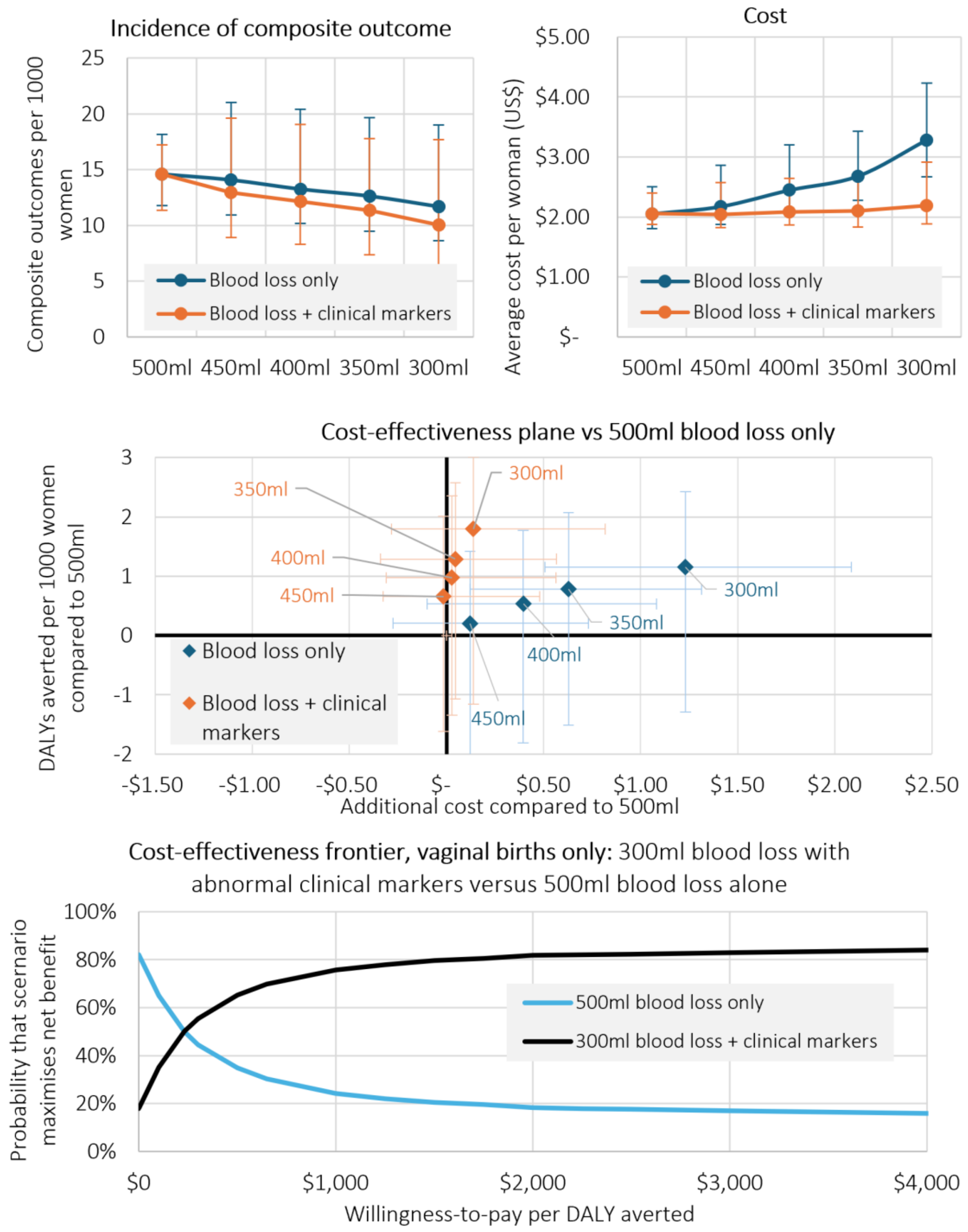


**Figure A1: Sub-analysis considering vaginal births only**. Model outcomes for blood loss thresholds alone ("blood loss only"), or blood loss thresholds in combination with any other abnormal haemodynamic marker ("blood loss + clinical markers"). Top left: incidence of composite outcome per 1000 women; top right: mean cost per woman (in 2022 US$); middle: disability-adjusted life years (DALYs) averted per 1000 women and difference in mean cost for scenarios compared to the 500 ml blood loss model; bottom: cost-effectiveness frontier, showing the probability that a scenario maximises net benefits for a given willingness-to-pay per DALY averted.

**Sensitivity analysis for high-risk populations**

When the incidence of the composite outcome was increased from 2.5% (as observed in the individual participant data) in the main analysis to 10%, then the impact of expanded treatment initiation criteria increased and additional costs were saved from composite outcomes averted. The lower blood loss thresholds were more cost-effective, and the lower blood loss thresholds in combination with any other abnormal haemodynamic marker were cost saving (Figure A2, Table A2). As per the main analysis, each scenario considering other abnormal haemodynamic markers remained more cost-effective than any of the blood loss only scenarios, due to reduced unnecessary treatment.

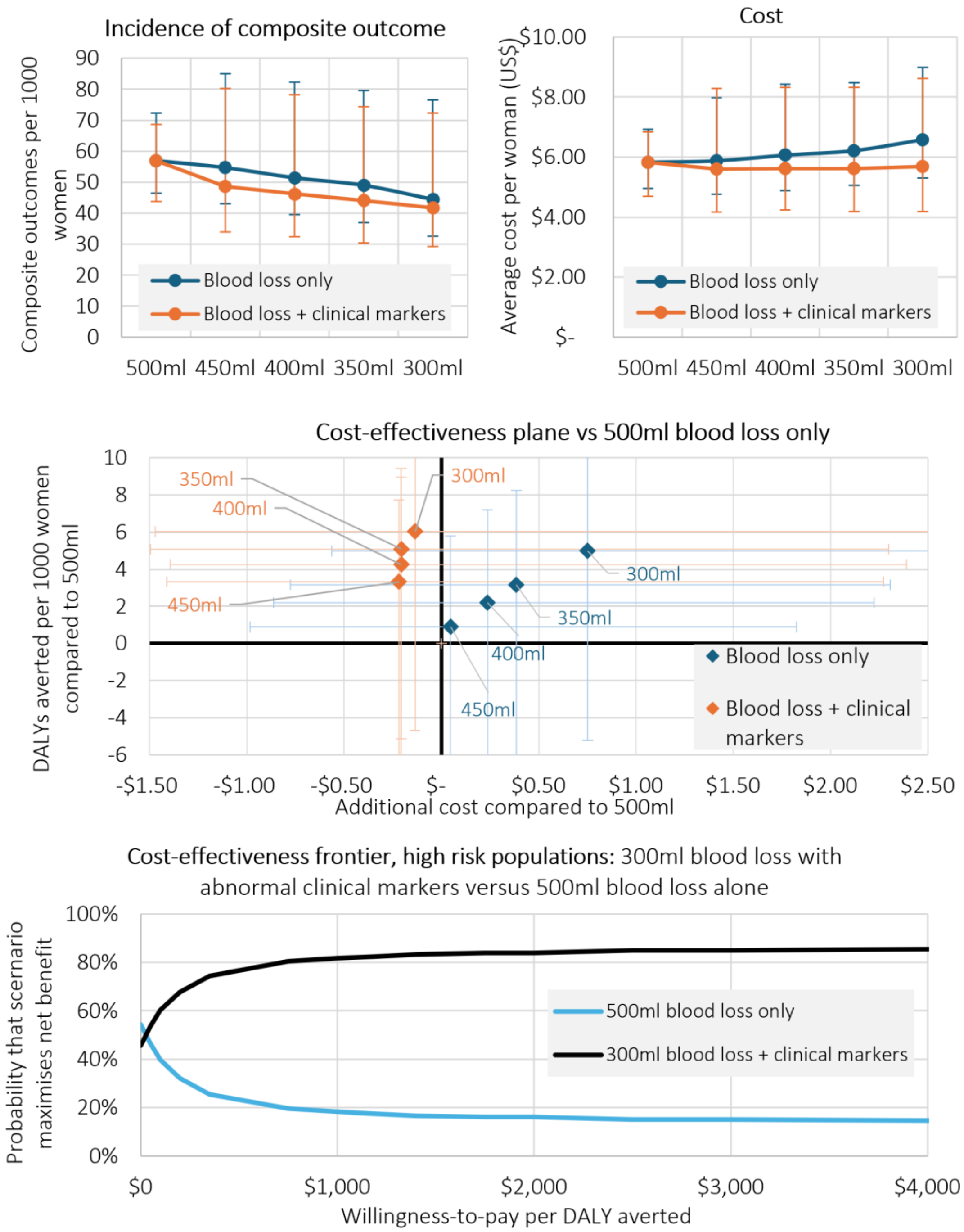


**Figure A2: Sub-analysis for higher risk populations (10% incidence of composite outcome compared to 2.5% in main analysis)**. Model outcomes for blood loss thresholds alone ("blood loss only"), or blood loss thresholds in combination with any other abnormal haemodynamic marker ("blood loss + clinical markers"). Top left: incidence of composite outcome per 1000 women; top right: mean cost per woman (in 2022 US$); middle: composite outcomes averted per 1000 women and difference in mean cost for scenarios compared to the 500 ml blood loss model; bottom: cost-effectiveness frontier, showing the probability that a scenario maximises net benefits for a given willingness-to-pay per composite outcome averted.

Table A2: Results for sensitivity analysis for higher risk populations (10% incidence of composite outcome compared to 2.5% in main analysis).

| Scenario | Incidence of composite outcome per 1000 women | Mean DALYs lost per 1000 women | Mean cost per woman (2022 US$) | Unnecessary treatments per 1000 women | Difference in composite outcomes per 1000 women, compared to 500 ml blood loss | Difference in DALYs per 1000 women, compared to 500 ml blood loss | Mean additional cost per woman compared to 500 ml blood loss (2022 US$) | Cost per DALY averted compared to 500 ml blood loss (2022 US$) |
|---|---|---|---|---|---|---|---|---|
| Blood loss | | | | | | | | |
| ≥ 500 ml blood loss | 57.0 (46.4, 72.3) | 22.5 (18.4, 28.7) | $5.83 ($4.96, $6.93) | 174 (117, 241) | | | | |
| ≥ 450 ml blood loss | 54.7 (43.0, 85.0) | 21.6 (17.1, 33.7) | $5.87 ($4.76, $7.98) | 205 (140, 280) | -2.3 (-14.7, 21.4) | -0.9 (-5.8, 8.5) | $0.05 (-$0.98, $1.82) | $51 (-$1576, $965) |
| ≥ 400 ml blood loss | 51.4 (39.5, 82.3) | 20.3 (15.7, 32.6) | $6.06 ($4.89, $8.43) | 267 (180, 366) | -5.6 (-18.1, 18.5) | -2.2 (-7.2, 7.3) | $0.24 (-$0.86, $2.22) | $107 (-$2394, $2956) |
| ≥ 350 ml blood loss | 49.0 (37.1, 79.6) | 19.3 (14.7, 31.6) | $6.21 ($5.06, $8.47) | 316 (216, 413) | -8.0 (-20.7, 15.9) | -3.2 (-8.2, 6.3) | $0.38 (-$0.78, $2.31) | $122 (-$3035, $2758) |
| ≥ 300 ml blood loss | 44.4 (32.6, 76.5) | 17.5 (12.9, 30.3) | $6.58 ($5.30, $8.98) | 409 (298, 529) | -12.6 (-25.9, 13.2) | -5.0 (-10.3, 5.2) | $0.75 (-$0.56, $2.78) | $151 (-$2963, $4383) |
| Blood loss + clinical marker(s) | | | | | | | | |
| ≥ 450 ml blood loss + clinical marker(s) | 48.6 (34.0, 80.2) | 19.2 (13.5, 31.7) | $5.61 ($4.17, $8.28) | 237 (131, 359) | -8.4 (-19.4, 18.7) | -3.3 (-7.7, 7.4) | -$0.22 (-$1.41, $2.27) | -$66 (-$2572, $1720) |
| ≥ 400 ml blood loss + clinical marker(s) | 46.2 (32.5, 78.3) | 18.2 (12.9, 31.0) | $5.62 ($4.25, $8.33) | 268 (147, 401) | -10.8 (-22.6, 16.4) | -4.3 (-8.9, 6.4) | -$0.21 (-$1.39, $2.39) | -$48 (-$3246, $2946) |
| ≥ 350 ml blood loss + clinical marker(s) | 44.1 (30.4, 74.4) | 17.4 (12.1, 29.4) | $5.62 ($4.18, $8.33) | 278 (156, 426) | -12.9 (-23.9, 13.0) | -5.1 (-9.4, 5.1) | -$0.21 (-$1.50, $2.30) | -$41 (-$2815, $2177) |
| ≥ 300 ml blood loss + clinical marker(s) | 41.7 (29.3, 72.3) | 16.5 (11.6, 28.6) | $5.69 ($4.19, $8.62) | 321 (184, 468) | -15.3 (-26.5, 11.9) | -6.0 (-10.5, 4.7) | -$0.14 (-$1.47, $2.59) | -$23 (-$2470, $2375) |

**Sensitivity analysis excluding effects of early treatment**

Without the early treatment effect, the impact of each scenario was reduced, and primarily a function of the scenario's sensitivity input parameters. The cost per composite outcome averted increased for all scenarios (Figure A3, Table A3).

As per the main analysis, each scenario considering other abnormal haemodynamic markers remained more cost-effective than any of the blood loss only scenarios, due to reduced unnecessary treatment.

When the threshold of 300 ml blood loss in combination with any other abnormal haemodynamic marker was compared to the threshold of 500 ml blood loss alone, it was the preferred strategy at a willingness-to-pay of >$861 per DALY averted.

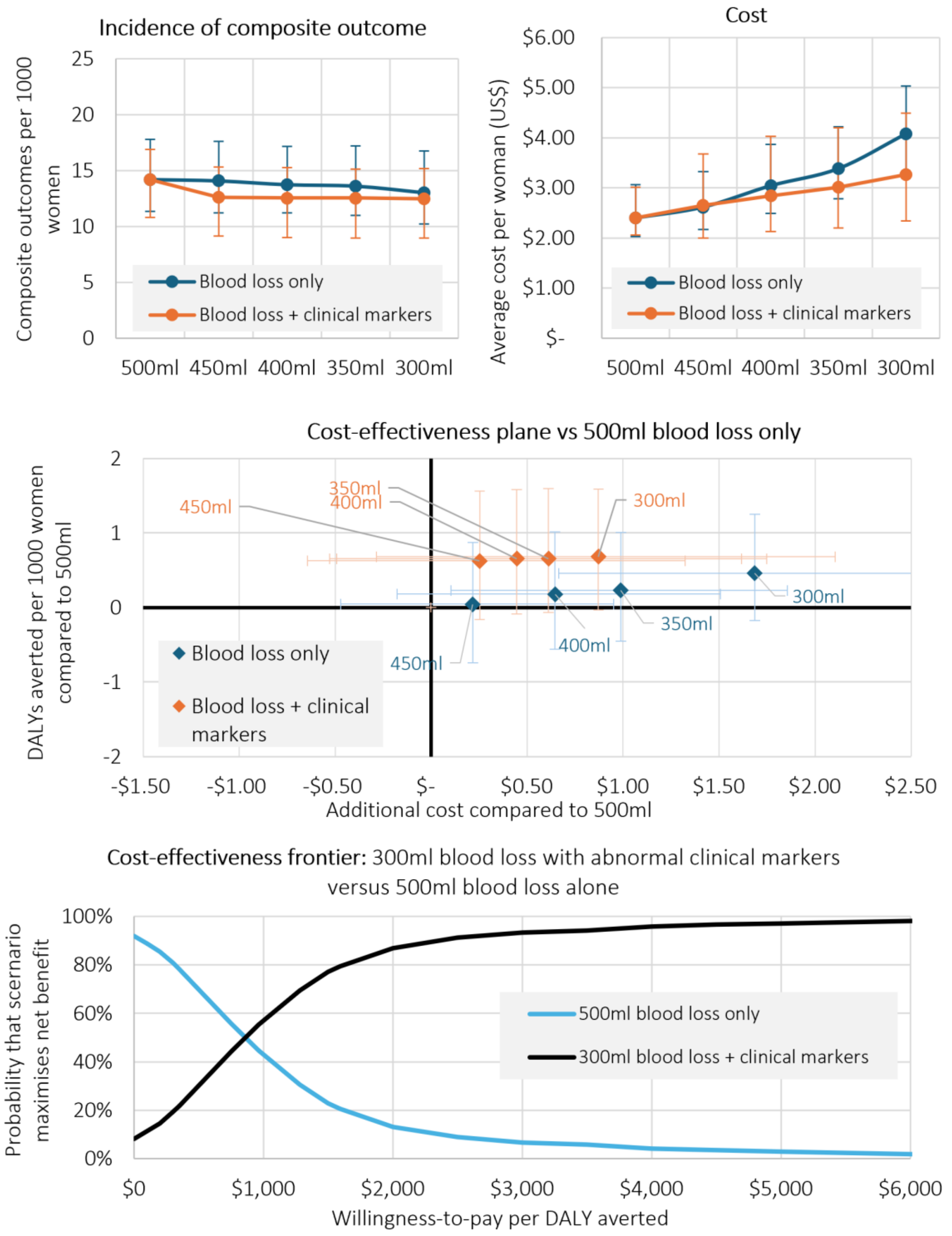


**Figure A3: Sensitivity analysis excluding effects of early treatment**. Model outcomes for blood loss thresholds alone ("blood loss only"), or blood loss thresholds in combination with any other abnormal haemodynamic marker ("blood loss + clinical markers"). Top left: incidence of composite outcome per 1000 women; top right: mean cost per woman (in 2022 US$); middle: composite outcomes averted per 1000 women and difference in mean cost for scenarios compared to the 500 ml blood loss model; bottom: cost-effectiveness frontier, showing the probability that a scenario maximises net benefits for a given willingness-to-pay per composite outcome averted.

Table A3: Results for sensitivity analysis excluding effects of early treatment.

| Scenario | Incidence of composite outcome per 1000 women | Mean DALYs lost per 1000 women | Mean cost per woman (2022 US$) | Unnecessary treatments per 1000 women | Difference in composite outcomes per 1000 women, compared to 500 ml blood loss | Difference in DALYs per 1000 women, compared to 500 ml blood loss | Mean additional cost per woman compared to 500 ml blood loss (2022 US$) | Cost per DALY averted compared to 500 ml blood loss (2022 US$) |
|---|---|---|---|---|---|---|---|---|
| Blood loss | | | | | | | | |
| ≥ 500 ml blood loss | 14.2 (11.4, 17.8) | 5.6 (4.5, 7.1) | $2.40 ($2.02, $3.06) | 189 (127, 261) | | | | |
| ≥ 450 ml blood loss | 14.1 (11.2, 17.6) | 5.6 (4.4, 7.0) | $2.62 ($2.17, $3.33) | 222 (152, 304) | -0.1 (-2.2, 1.9) | 0.0 (-0.9, 0.7) | $0.22 (-$0.47, $0.95) | $4872 (-$14892, $15711) |
| ≥ 400 ml blood loss | 13.7 (11.2, 17.2) | 5.4 (4.4, 6.8) | $3.04 ($2.49, $3.87) | 289 (196, 396) | -0.5 (-2.5, 1.4) | -0.2 (-1.0, 0.6) | $0.65 (-$0.18, $1.51) | $3615 (-$26983, $30478) |
| ≥ 350 ml blood loss | 13.6 (11.0, 17.2) | 5.4 (4.4, 6.8) | $3.39 ($2.78, $4.22) | 342 (235, 448) | -0.6 (-2.5, 1.1) | -0.2 (-1.0, 0.4) | $0.99 ($0.10, $1.86) | $4316 (-$29984, $39064) |
| ≥ 300 ml blood loss | 13.0 (10.2, 16.7) | 5.1 (4.1, 6.6) | $4.08 ($3.29, $5.03) | 444 (324, 574) | -1.2 (-3.1, 0.4) | -0.5 (-1.3, 0.2) | $1.69 ($0.66, $2.58) | $3682 (-$28407, $32645) |
| Blood loss + clinical marker(s) | | | | | | | | |
| ≥ 450 ml blood loss + clinical marker(s) | 12.6 (9.1, 15.3) | 5.0 (3.6, 6.1) | $2.65 ($2.00, $3.68) | 253 (142, 387) | -1.6 (-4.0, 0.4) | -0.6 (-1.6, 0.2) | $0.25 (-$0.65, $1.32) | $404 (-$3130, $5654) |
| ≥ 400 ml blood loss + clinical marker(s) | 12.5 (9.0, 15.3) | 5.0 (3.6, 6.0) | $2.84 ($2.13, $4.03) | 287 (160, 433) | -1.7 (-4.0, 0.2) | -0.7 (-1.6, 0.1) | $0.45 (-$0.53, $1.62) | $684 (-$2112, $6126) |
| ≥ 350 ml blood loss + clinical marker(s) | 12.5 (9.0, 15.1) | 5.0 (3.6, 6.0) | $3.01 ($2.20, $4.20) | 298 (169, 459) | -1.7 (-4.0, 0.2) | -0.7 (-1.6, 0.1) | $0.61 (-$0.49, $1.75) | $936 (-$4369, $7957) |
| ≥ 300 ml blood loss + clinical marker(s) | 12.5 (8.9, 15.2) | 4.9 (3.6, 6.0) | $3.27 ($2.34, $4.49) | 345 (198, 506) | -1.7 (-4.0, 0.1) | -0.7 (-1.6, 0.0) | $0.87 (-$0.29, $2.11) | $1279 (-$3865, $10222) |

#### APPENDIX B: Sensitivity analysis, treatment and composite outcome unit costs

*Appendix B_unit cost sensitivity analysis.xlsx*

#### APPENDIX C: Additional outputs

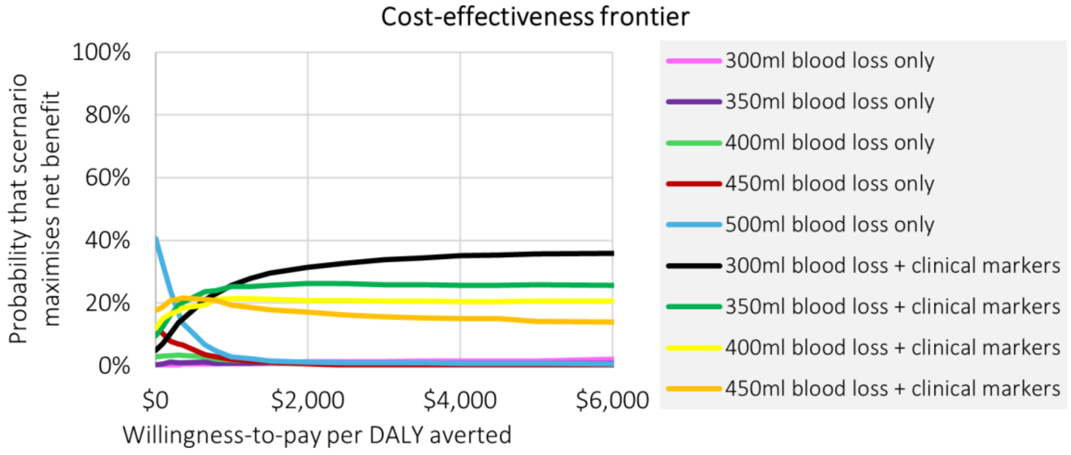


**Figure C1: Cost-effectiveness frontier comparing all scenarios.** Figure shows the probability that a scenario maximises net benefits for a given willingness-to-pay per composite outcome averted. Blood loss only – model outcomes for blood loss thresholds alone; blood loss + clinical markers – model outcomes for blood loss thresholds in combination with any other abnormal haemodynamic marker.

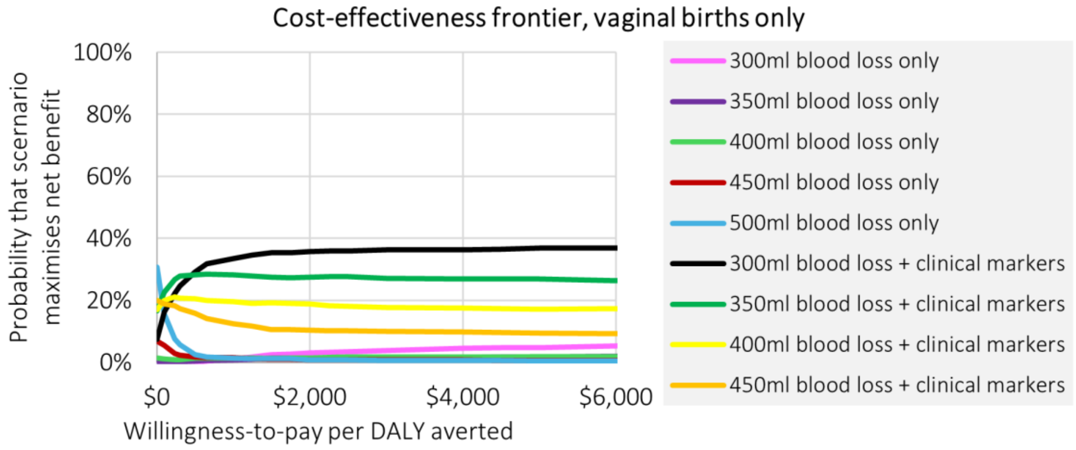


**Figure C2: Cost-effectiveness frontier comparing all scenarios, vaginal births only.** Figure shows the probability that a scenario maximises net benefits for a given willingness-to-pay per composite outcome averted. Blood loss only – model outcomes for blood loss thresholds alone; blood loss + clinical markers – model outcomes for blood loss thresholds in combination with any other abnormal haemodynamic marker.

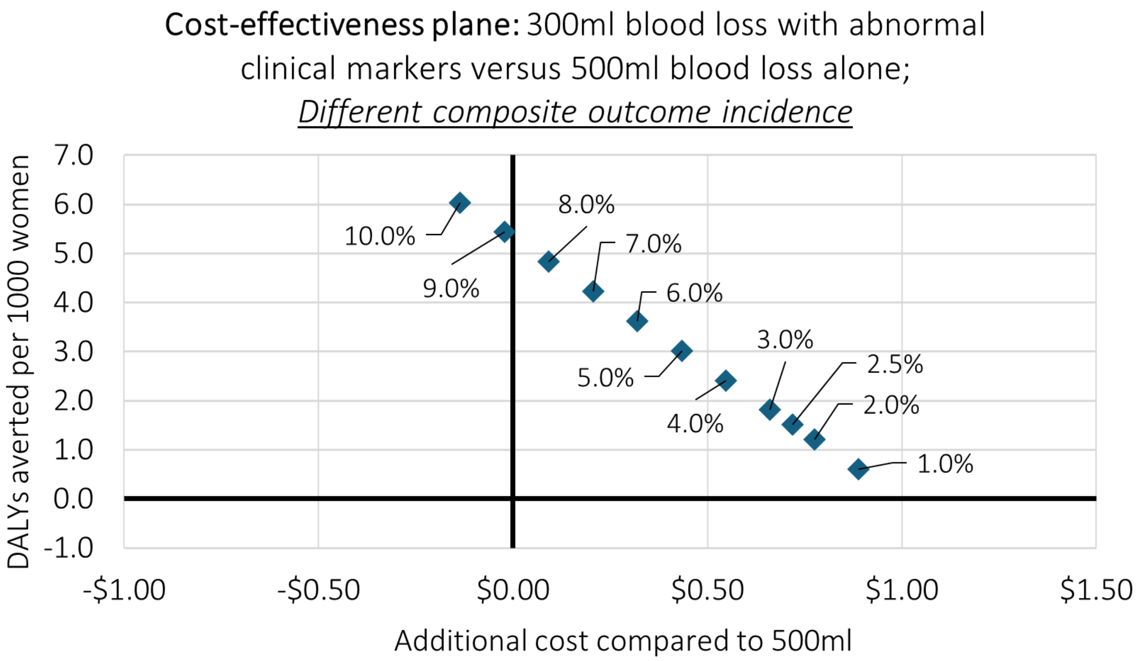


**Figure C3: Cost-effectiveness plane for 300 ml blood loss + clinical markers compared to 500 ml blood loss only, <u>different incidence of composite outcome</u>.**

Table C1: Cost per DALY averted as a percentage of GDP per capita for low- and middle-income countries. The 300 ml blood loss in combination with any other abnormal haemodynamic marker compared to the 500 ml blood loss alone.

| | | | Scenario, compared to 500 ml blood loss alone | 300 ml blood loss + clinical marker(s) (main analysis) | 300 ml blood loss + clinical marker(s) (vaginal births only) |
|---|---|---|---|---|---|
| | | | ICER for scenario | $479 | $77 |
| Country ISO3 | Country name | Income status | GDP per capita in 2022 (2022 US$) | ICER as a percentage of GDP per capita (main analysis) | ICER as a percentage of GDP per capita (vaginal births only) |
| Afghanistan | AFG | Low income | $357 | 134% | 22% |
| Angola | AGO | Lower middle income | $2,930 | 16% | 3% |
| Albania | ALB | Upper middle income | $6,846 | 7% | 1% |
| Argentina | ARG | Upper middle income | $13,936 | 3% | 1% |
| Armenia | ARM | Upper middle income | $6,572 | 7% | 1% |
| Azerbaijan | AZE | Upper middle income | $7,771 | 6% | 1% |
| Burundi | BDI | Low income | $251 | 191% | 31% |
| Benin | BEN | Lower middle income | $1,265 | 38% | 6% |
| Burkina Faso | BFA | Low income | $836 | 57% | 9% |
| Bangladesh | BGD | Lower middle income | $2,716 | 18% | 3% |

| | | | | | |
|---|---|---|---|---|---|
| Bosnia and Herzegovina | BIH | Upper middle income | $7,656 | 6% | 1% |
| Belarus | BLR | Upper middle income | $7,995 | 6% | 1% |
| Belize | BLZ | Upper middle income | $7,068 | 7% | 1% |
| Bolivia | BOL | Lower middle income | $3,644 | 13% | 2% |
| Brazil | BRA | Upper middle income | $9,281 | 5% | 1% |
| Botswana | BWA | Upper middle income | $8,329 | 6% | 1% |
| Central African Republic | CAF | Low income | $467 | 102% | 16% |
| China | CHN | Upper middle income | $12,663 | 4% | 1% |
| Cote d'Ivoire | CIV | Lower middle income | $2,309 | 21% | 3% |
| Cameroon | CMR | Lower middle income | $1,605 | 30% | 5% |
| Congo, Dem. Rep. | COD | Low income | $643 | 74% | 12% |
| Congo, Rep. | COG | Lower middle income | $2,621 | 18% | 3% |
| Colombia | COL | Upper middle income | $6,675 | 7% | 1% |
| Comoros | COM | Lower middle income | $1,489 | 32% | 5% |
| Cabo Verde | CPV | Lower middle income | $4,323 | 10% | 2% |
| Costa Rica | CRI | Upper middle income | $13,626 | 3% | 0% |
| Djibouti | DJI | Lower middle income | $3,231 | 13% | 2% |
| Dominica | DMA | Upper middle income | $9,086 | 5% | 1% |
| Dominican Republic | DOM | Upper middle income | $10,110 | 4% | 1% |
| Algeria | DZA | Upper middle income | $4,962 | 9% | 1% |
| Ecuador | ECU | Upper middle income | $6,541 | 7% | 1% |
| Egypt, Arab Rep. | EGY | Lower middle income | $4,233 | 14% | 2% |
| Ethiopia | ETH | Low income | $1,011 | 38% | 6% |
| Fiji | FJI | Upper middle income | $5,405 | 8% | 1% |
| Micronesia, Fed. Sts. | FSM | Lower middle income | $3,835 | 12% | 2% |
| Gabon | GAB | Upper middle income | $8,409 | 6% | 1% |
| Georgia | GEO | Upper middle income | $6,730 | 6% | 1% |
| Ghana | GHA | Lower middle income | $2,240 | 21% | 3% |
| Guinea | GIN | Lower middle income | $1,417 | 31% | 5% |
| Gambia, The | GMB | Low income | $836 | 54% | 9% |
| Guinea-Bissau | GNB | Low income | $853 | 50% | 8% |
| Equatorial Guinea | GNQ | Upper middle income | $7,589 | 7% | 1% |
| Grenada | GRD | Upper middle income | $10,474 | 4% | 1% |
| Guatemala | GTM | Upper middle income | $5,358 | 8% | 1% |
| Honduras | HND | Lower middle income | $3,003 | 15% | 2% |
| Haiti | HTI | Lower middle income | $1,761 | 28% | 5% |
| Indonesia | IDN | Upper middle income | $4,731 | 10% | 2% |
| India | IND | Lower middle income | $2,353 | 19% | 3% |
| Iran, Islamic Rep. | IRN | Upper middle income | $4,405 | 11% | 2% |
| Iraq | IRQ | Upper middle income | $6,504 | 9% | 1% |
| Jamaica | JAM | Upper middle income | $6,022 | 7% | 1% |
| Jordan | JOR | Lower middle income | $4,332 | 11% | 2% |
| Kazakhstan | KAZ | Upper middle income | $11,255 | 4% | 1% |
| Kenya | KEN | Lower middle income | $2,110 | 25% | 4% |
| Kyrgyz Republic | KGZ | Lower middle income | $1,740 | 24% | 4% |
| Cambodia | KHM | Lower middle income | $2,325 | 20% | 3% |
| Kiribati | KIR | Lower middle income | $2,076 | 23% | 4% |
| Lao PDR | LAO | Lower middle income | $2,046 | 23% | 4% |
| Liberia | LBR | Low income | $745 | 62% | 10% |
| Libya | LBY | Upper middle income | $5,987 | 8% | 1% |
| St. Lucia | LCA | Upper middle income | $13,104 | 4% | 1% |

| | | | | | |
|---|---|---|---|---|---|
| Sri Lanka | LKA | Lower middle income | $3,343 | 13% | 2% |
| Lesotho | LSO | Lower middle income | $1,030 | 52% | 8% |
| Morocco | MAR | Lower middle income | $3,455 | 13% | 2% |
| Moldova | MDA | Upper middle income | $5,738 | 7% | 1% |
| Madagascar | MDG | Low income | $497 | 95% | 15% |
| Maldives | MDV | Upper middle income | $11,786 | 4% | 1% |
| Mexico | MEX | Upper middle income | $11,385 | 3% | 1% |
| Marshall Islands | MHL | Upper middle income | $6,323 | 7% | 1% |
| North Macedonia | MKD | Upper middle income | $7,606 | 6% | 1% |
| Mali | MLI | Low income | $814 | 55% | 9% |
| Myanmar | MMR | Lower middle income | $1,158 | 39% | 6% |
| Montenegro | MNE | Upper middle income | $10,093 | 4% | 1% |
| Mongolia | MNG | Upper middle income | $4,994 | 8% | 1% |
| Mozambique | MOZ | Low income | $578 | 77% | 12% |
| Mauritania | MRT | Lower middle income | $1,960 | 23% | 4% |
| Mauritius | MUS | Upper middle income | $10,240 | 4% | 1% |
| Malawi | MWI | Low income | $604 | 79% | 13% |
| Malaysia | MYS | Upper middle income | $11,748 | 4% | 1% |
| Namibia | NAM | Upper middle income | $4,349 | 11% | 2% |
| Niger | NER | Low income | $610 | 74% | 12% |
| Nigeria | NGA | Lower middle income | $2,139 | 30% | 5% |
| Nicaragua | NIC | Lower middle income | $2,325 | 18% | 3% |
| Nepal | NPL | Lower middle income | $1,386 | 35% | 6% |
| Pakistan | PAK | Lower middle income | $1,538 | 35% | 6% |
| Peru | PER | Upper middle income | $7,363 | 6% | 1% |
| Philippines | PHL | Lower middle income | $3,548 | 13% | 2% |
| Papua New Guinea | PNG | Lower middle income | $3,102 | 16% | 3% |
| Paraguay | PRY | Upper middle income | $6,206 | 8% | 1% |
| West Bank and Gaza | PSE | Lower middle income | $3,800 | 14% | 2% |
| Rwanda | RWA | Low income | $975 | 47% | 8% |
| Sudan | SDN | Low income | $1,046 | 22% | 4% |
| Senegal | SEN | Lower middle income | $1,565 | 28% | 5% |
| Solomon Islands | SLB | Lower middle income | $2,005 | 23% | 4% |
| Sierra Leone | SLE | Low income | $860 | 63% | 10% |
| El Salvador | SLV | Upper middle income | $5,094 | 9% | 1% |
| Somalia | SOM | Low income | $573 | 80% | 13% |
| Serbia | SRB | Upper middle income | $10,023 | 4% | 1% |
| Sao Tome and Principe | STP | Lower middle income | $2,390 | 16% | 3% |
| Suriname | SUR | Upper middle income | $6,084 | 9% | 1% |
| Eswatini | SWZ | Lower middle income | $3,852 | 13% | 2% |
| Chad | TCD | Low income | $672 | 70% | 11% |
| Togo | TGO | Low income | $899 | 49% | 8% |
| Thailand | THA | Upper middle income | $6,909 | 7% | 1% |
| Tajikistan | TJK | Lower middle income | $1,052 | 41% | 7% |
| Turkmenistan | TKM | Upper middle income | $8,156 | 6% | 1% |
| Timor-Leste | TLS | Lower middle income | $2,343 | 32% | 5% |
| Tunisia | TUN | Lower middle income | $3,678 | 12% | 2% |
| Turkiye | TUR | Upper middle income | $10,675 | 4% | 1% |
| Tuvalu | TUV | Upper middle income | $5,911 | 8% | 1% |
| Tanzania | TZA | Lower middle income | $1,208 | 39% | 6% |
| Uganda | UGA | Low income | $963 | 48% | 8% |
| Ukraine | UKR | Upper middle income | $4,200 | 9% | 2% |

| | | | | | |
|---|---|---|---|---|---|
| Uzbekistan | UZB | Lower middle income | $2,579 | 17% | 3% |
| St. Vincent and the Grenadines | VCT | Upper middle income | $9,471 | 5% | 1% |
| Viet Nam | VNM | Lower middle income | $4,116 | 11% | 2% |
| Vanuatu | VUT | Lower middle income | $3,265 | 14% | 2% |
| Samoa | WSM | Lower middle income | $3,869 | 11% | 2% |
| Kosovo | XKX | Upper middle income | $5,291 | 8% | 1% |
| Yemen, Rep. | YEM | Low income | $616 | 100% | 16% |
| South Africa | ZAF | Upper middle income | $6,523 | 8% | 1% |
| Zambia | ZMB | Lower middle income | $1,447 | 36% | 6% |
| Zimbabwe | ZWE | Lower middle income | $2,041 | 22% | 4% |
